# Video Observed Therapy (VOT) for People with Tuberculosis (TB): A Scoping Review

**DOI:** 10.1101/2024.04.23.24306229

**Authors:** An Du Thinh, Eleanor Morgan, Kassia Pereira, Celso Khosa, Tom Wingfield

**Author notes:** Corresponding Author: Dr Tom Wingfield, X: @drtomwingfield, Room W-01-34 Wolfson Building, Liverpool School of Tropical Medicine, Liverpool, L3 5QA, United Kingdom. Contributed equally (Celso Khosa, Tom Wingfield).

## Abstract

**Background:** Tuberculosis (TB) remains a significant cause of morbidity and mortality globally, disproportionately affecting low- and middle-income countries (LMIC). Accessing Directly Observed Therapy (DOT) is associated with out-of-pocket costs and stigma. Video-observed therapy (VOT) is an alternative to DOT but evidence from LMIC with high TB burden is limited. To make recommendations for future VOT-related research and inform the design of a pilot implementation of VOT for people with multi-drug resistant TB (MDR-TB) in Mozambique, we did a scoping review of the evidence on VOT for people with TB.

**Methodology:** We systematically searched five scientific databases and key grey literature repositories to identify eligible abstracts. Abstracts were reviewed and full-text records were identified and evaluated. Data from full-text records were extracted into four implementation theme categories: Feasibility, Acceptability, Cost, and Effectiveness (FACE). Content analysis was used to describe implementation successes and challenges, comparing VOT versus DOT where possible. The Crowe Critical Appraisal Tool (CCAT) was used to evaluate the quality of studies.

**Results:** In total, 66 records were identified: 47 primary research studies, 13 reviews, and six grey literature documents. All studies were graded as moderate to high quality and reported against at least one FACE category. Studies from urban settings (n=34) and HIC (n=33) predominated. Where measured and described, VOT implementation was reported to be feasible (43/43, 100%) and acceptable (43/44, 97%). Of the 18/20 (90%) studies describing cost data, VOT was reported to offer savings to the health system compared to DOT. Patient costs were under-reported. Of the 21/23 (91%) studies describing effectiveness to improve adherence to TB treatment, VOT was reported to be non-inferior to DOT.

**Conclusion:** In HIC settings, VOT was reported as feasible, acceptable, and similarly effective alternative to DOT. Further evidence on VOT is needed from LMIC with high TB burden.

**Author Summary:** Directly Observed Therapy (DOT) remains widely used to monitor treatment adherence of people living with tuberculosis (TB). Since COVID-19, there has been a massive increase in digital health technologies, mobile phone ownership, and internet connectivity, including in high TB burden low- and middle-income countries (LMIC), highlighting the potential for Video Observed Therapy (VOT) as a suitable alternative to DOT. We did a scoping review of studies reporting the implementation and evaluation of VOT for people with TB using a novel thematic framework consisting of feasibility, acceptability, cost and effectiveness (FACE). We identified 20 VOT applications (45% freely available) used by studies. Most VOT studies were conducted in urban settings of low TB burden, High Income Countries (HIC), and did not include cost-effectiveness analyses from a patient perspective or participants from key, underserved groups such as pregnant women or people with TB/HIV. The available evidence suggested that, in mainly HIC with low TB burden, VOT was a feasible, acceptable and effective alternative to DOT which could provide cost savings to the healthcare provider. There is an urgent need for robust evidence of effectiveness, cost-effectiveness, and equity of VOT in LMICs with high TB burden and among key, underserved populations.

## Introduction

Tuberculosis (TB) remains a major global public health challenge. In 2023, TB regained, from COVID-19, the position of the leading cause of death from a single infectious disease worldwide^1,2^. Most high TB burden countries are lower- and lower-middle-income countries (LMIC), which are disproportionately affected by TB-related morbidity and mortality, especially where HIV prevalence is high^2,3^. Without treatment, 10-year mortality from TB is approximately 70%^3^. Although shorter TB treatment regimens are effective in treating TB, most countries are still using antimicrobial regimens lasting up to 18 months to treat multi-drug resistant TB (MDR-TB), a leading cause of death from antimicrobial resistance (AMR). The high pill-burden and long treatment duration has clinical, psychological, and socio-economic consequences, challenging adherence. Treatment success rates for MDR-TB are estimated to be approximately 60%^3^. Suboptimal adherence increases the likelihood of adverse TB treatment outcomes including treatment failure, AMR, and onward transmission.

Providing support to people with TB to facilitate adherence to treatment can minimize adverse outcomes^4^. Directly Observed Therapy (DOT) has been recommended by the World Health Organization (WHO) as a method to improve TB treatment adherence. While some studies have suggested that DOT can increase adherence when compared to self-administered therapy (SAT), a Cochrane systematic review and meta-analysis found no significant differences in treatment success between DOT and SAT^4^. Facility-based DOT involves observation of medication doses by healthcare workers (HCW) at healthcare centres. This requires daily travel by people with TB and DOT may not be accessible for communities in remote locations, where travel to healthcare centres is challenging or unaffordable^5^. In many cases this results in high levels of SAT, especially during weekends when healthcare centres may be closed. Moreover, DOT may be associated with stigma and a loss of autonomy among people with TB^6^.

Video Observed therapy (VOT) is a potential alternative to DOT. In VOT, videos are live-streamed or uploaded for review by HCWs, either *synchronously* (at the time of medication ingestion) or *asynchronously* (the recording is reviewed after medication ingestion). VOT has been highlighted as an essential digital health tool to achieve the 2015 WHO End TB Strategy aims to eliminate TB^7^. In some studies^8,9^, VOT has shown promise in increasing treatment adherence. This may relate to VOT being more convenient than DOT because it allows people with TB to take their treatment from home, work, or another location, using their mobile device to record ingestion. Moreover, compared to an in-person DOT appointment at a healthcare centre, VOT can reduce travel time, health system costs, patient costs, and patients’ perceived stigma^7,10,11^.

Despite its potential, there is limited up-to-date evidence on the implementation and effectiveness of VOT in LMIC and amongst vulnerable groups, including people with MDR-TB or comorbidities. This evidence gap is particularly pertinent given the expansion of digital health technologies since the COVID-19 pandemic and the increase in mobile phone ownership, usage, and internet coverage in LMICs^12^. This scoping review aimed to gather, synthesise and critically appraise the latest evidence on VOT for people with TB globally in order to inform the design of a pilot implementation of VOT for people with MDR-TB in Mozambique (the “SAFEST-1 MDR-TB” study), and make recommendations for future research.

## Methods

### Study Design

A scoping review was considered the most suitable method to comprehensively gather evidence from both the scientific and grey literature, provide details on VOT platforms used, and encompass heterogeneous study types and resources^13,14^. The scoping review was conducted in accordance with the Joanna Briggs Institute (JBI) guidance^14^.

The JBI and Cochrane recommended PICO (Population, Intervention, Comparison, and Outcome) was used to help inform the search strategy and inclusion criteria^15^. Population was adults, pregnant women, adolescents, and children with TB. Intervention was VOT for TB treatment adherence monitoring. Comparison was DOT, DOTS, or SAT vs VOT. Outcomes included feasibility, acceptability, costs, effectiveness, and reported success and challenges of implementation.

### Inclusion Criteria

The inclusion criteria included: i) Accessible peer-reviewed scientific journal articles of any study design or published reports with the primary or co-primary aim of describing and/or evaluating and/or reviewing Video Observed Therapy (VOT) for people (adults and children) with tuberculosis (TB) of any type and in any country, and assessed to be of sufficient quality to facilitate suitable data extraction for meaningful content analysis and interpretation (detailed further below); ii) Reports, working papers, technical guidance, and other non-scientific literature appearing in the grey literature of key databases (detailed further below); iii) Publication between 1^st^ of January 2000 and 1^st^ of January 2023 – a timescale chosen because of the paucity of digital adherence technologies, video-phone applications, and mobile phone coverage prior to the year 2000; and iv) Publications in English, Portuguese, French, and Spanish languages, in which the research team collectively had suitable reading comprehension skills.

### Search Strategy

We systematically searched Medline, Embase, PubMed, Google Scholar and Web of Science databases for relevant articles, supplemented by a grey literature search of key databases to identify records published between 1st January 2000 and 1st January 2023. Grey literature was identified from a pre-specified convenience sample of databases selected through the consensus of the research team. These included: the World Health Organisation (WHO), Centers for Disease Control and Prevention (CDC), European Centre for Disease Prevention and Control (ECDC), and WHO International Clinical Trials Registry Platform (ICTRP). Reference snowballing and citation tracking were used to complement and expand the search.

The search for records was conducted between 9th February 2023 and 31st March 2023. Duplicates were identified and removed. Titles and abstracts were reviewed by two team members independently, but not blinded, according to the inclusion criteria. Where there was disagreement, a third opinion was sought from a third reviewer. The full-text of selected records were uploaded to a password-protected online, “live” shared Excel spreadsheet stored on an LSTM cloud-based server. Only deidentified and anonymised secondary data was stored and used and there was no confidential participant information.

The quality of records selected for full-text review was evaluated using the Crowe Critical Appraisal Tool (CCAT)^16^. Based on previous research by our study team^6^, a decision was taken to pragmatically categorise CCAT scores (percentage score out of a total of 40 points) as follows: ≥30 points, 75%-100%, high quality; 20 to 29 points, 50%-74%, moderate quality; and <20 points, <50%, low quality.

### Data Analysis

From included full-text records, we extracted aggregate sociodemographic data of participants, geographic and World Bank economic classification data (low, middle, high income) of study sites, data related to the VOT platforms used, and implementation data for content analysis on the challenges and success of VOT with relation to Feasibility, Acceptability, Cost, and Effectiveness (FACE) – categories decided a priori through study team consensus. Data were presented in narrative and tabular form.

## Results

### Study Selection

The search yielded 429 records: PubMed (n=138), Medline (n=79), Web of Science (n=100), Cochrane (n=7) and Google Scholar (n=105)(Figure 1). Of these, 160 were removed, mainly due to duplication, leaving 269 records to be screened. Following eligibility screening, 60 records were selected for full-text review. The complementary search of the pre-specified convenience sample of grey literature databases yielded 22 registered clinical trials and citation “snowballing” searching by manually reviewing references of the above 60 included studies found another six relevant studies; these two sources combined to identify a further six studies for full-text review (Figure 1).

**Figure 1:**
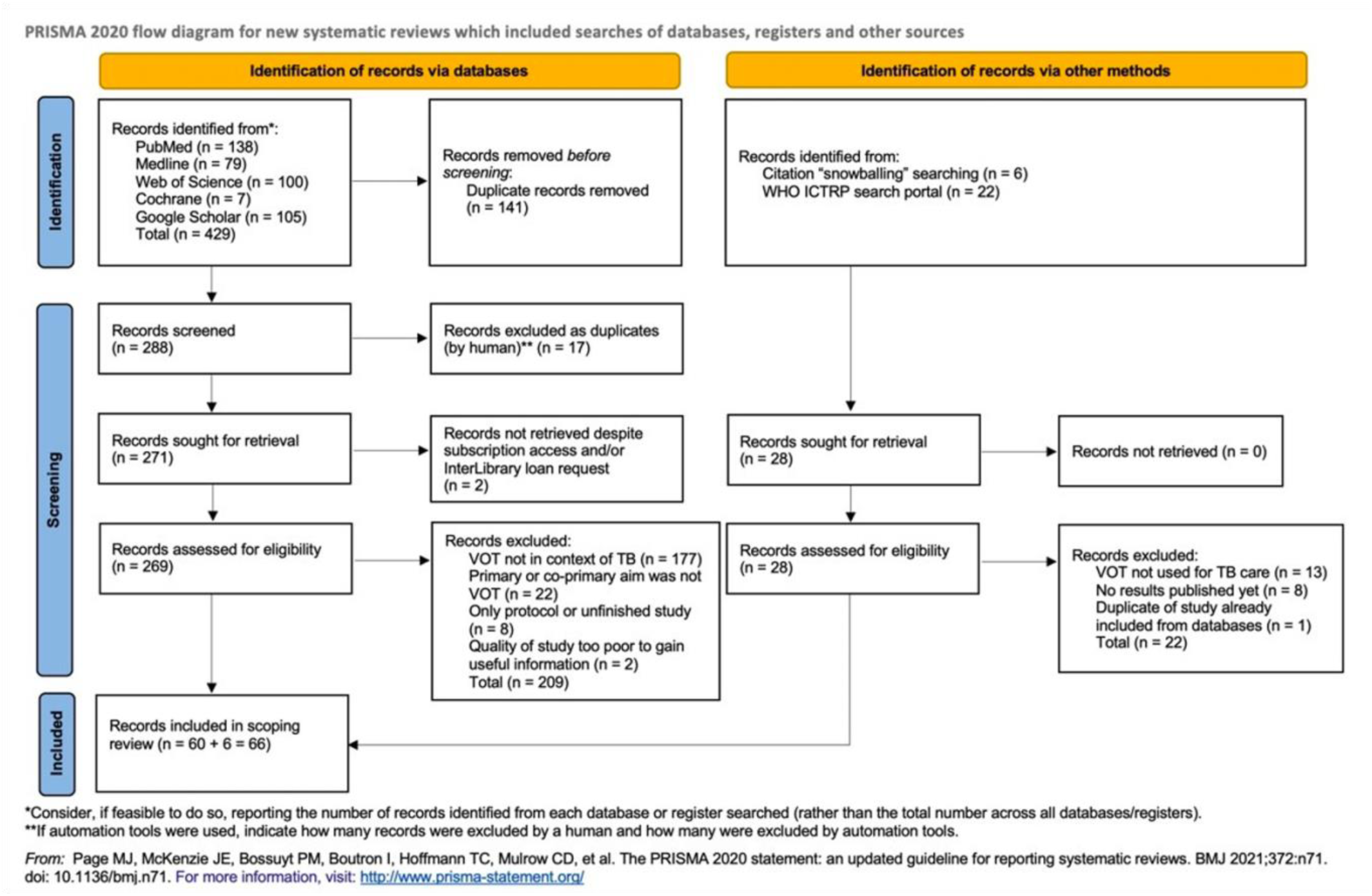
PRISMA-ScR flow diagram.

### Study Characteristics

The 66 records selected consisted of: 47 primary research studies (Table 1), 13 review-type studies including systematic reviews, scoping reviews, and narrative/non-systematic reviews (Table 2), and six grey literature documents (Table 3).

**Table 1:**
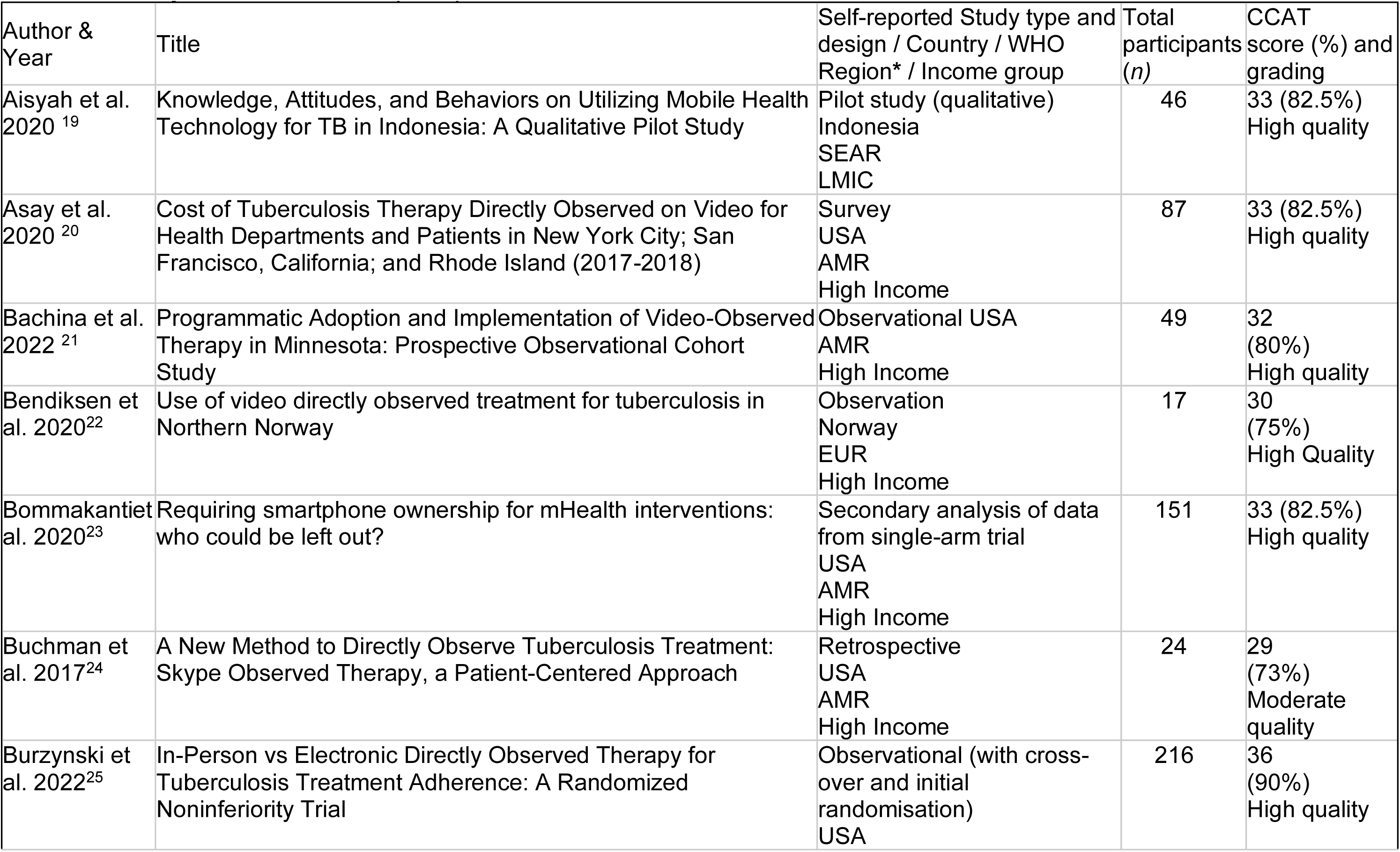
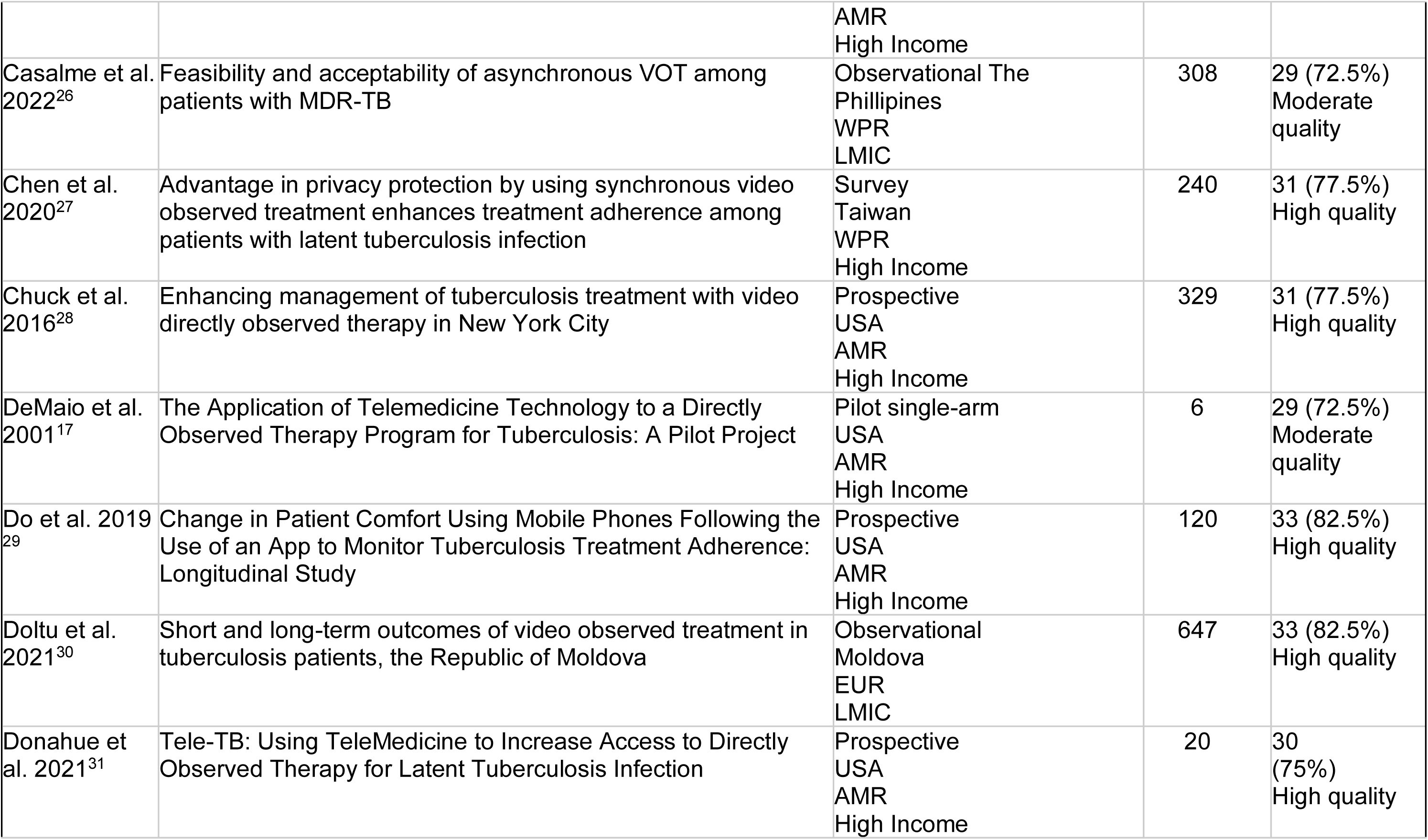
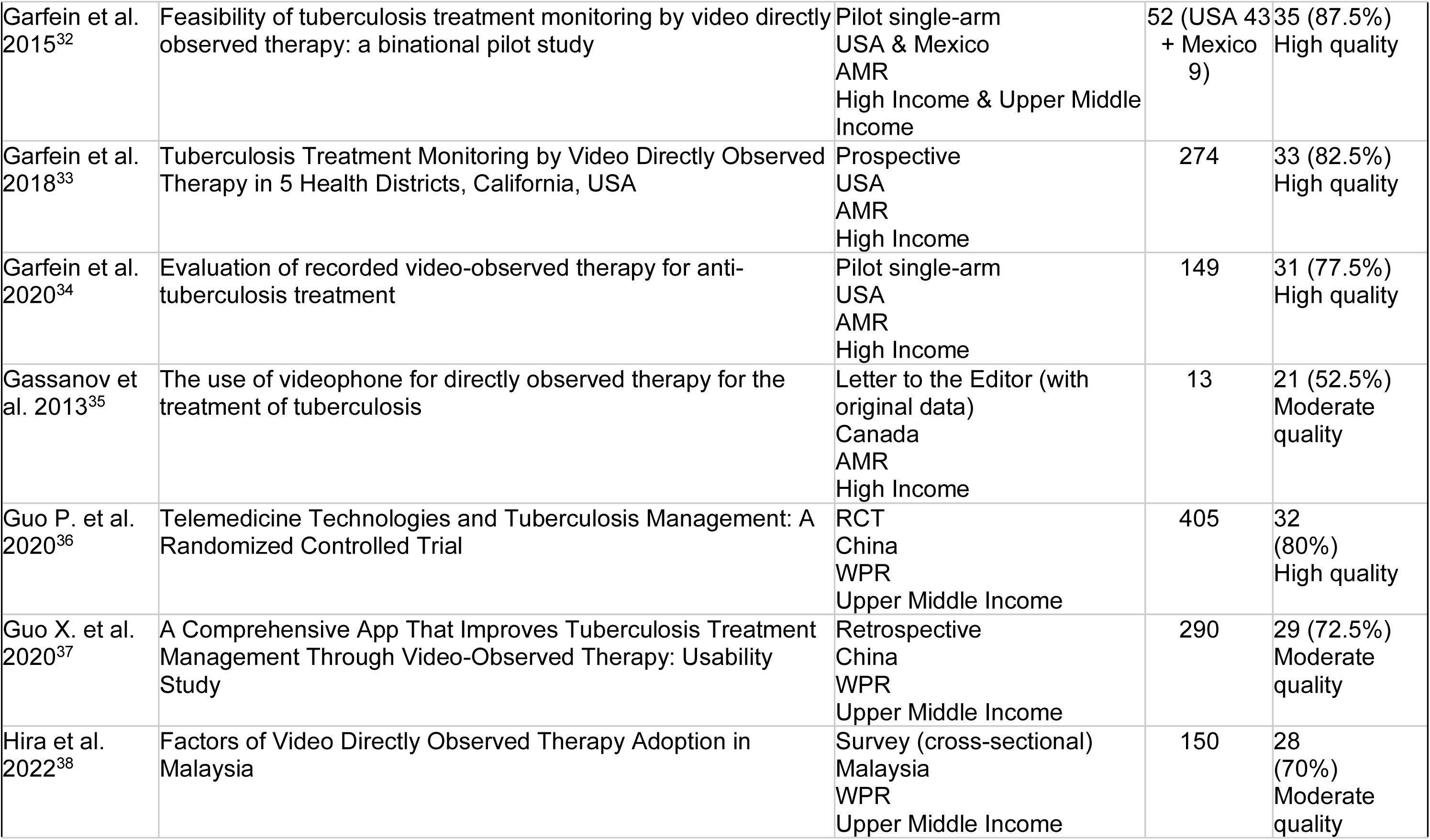
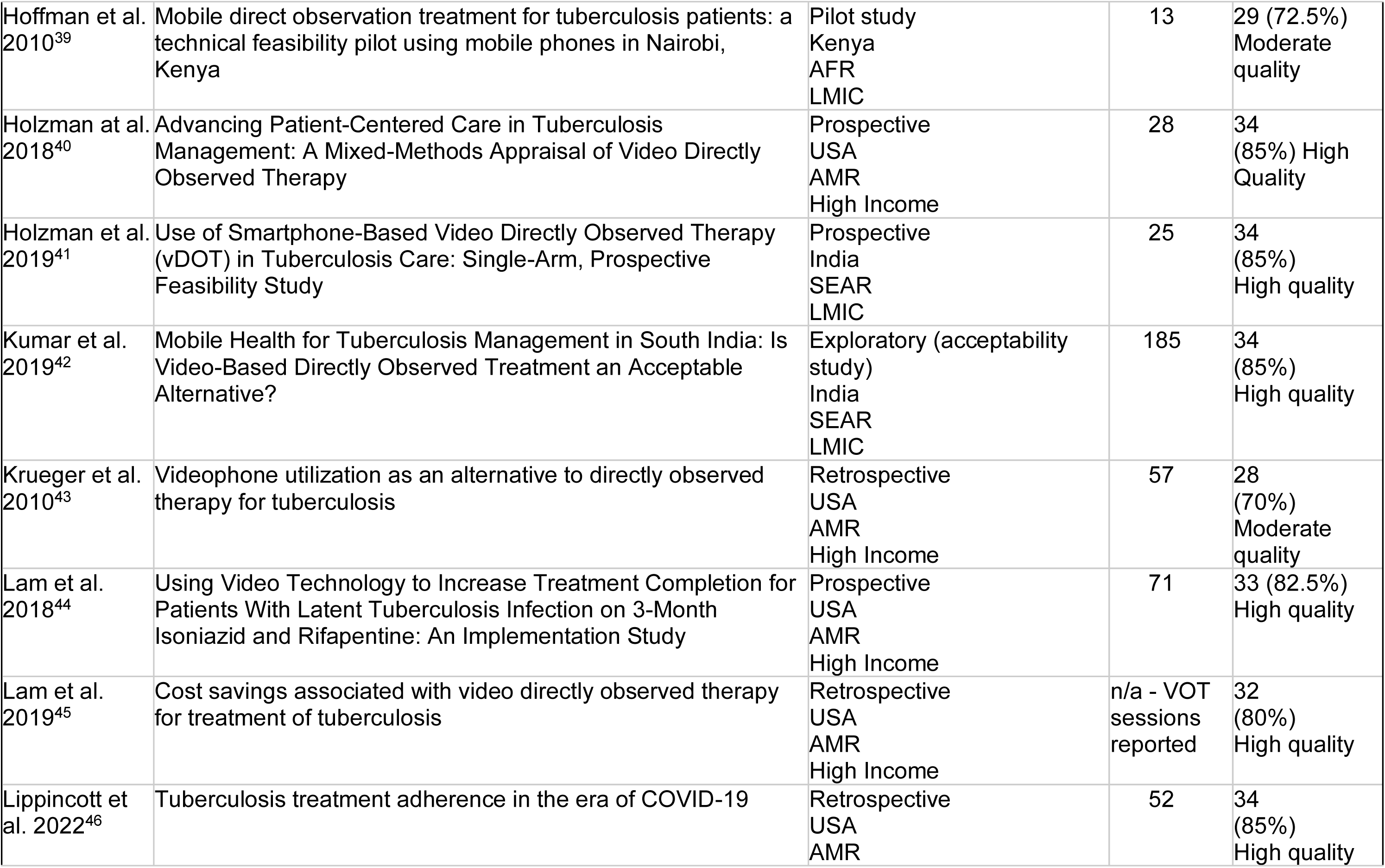
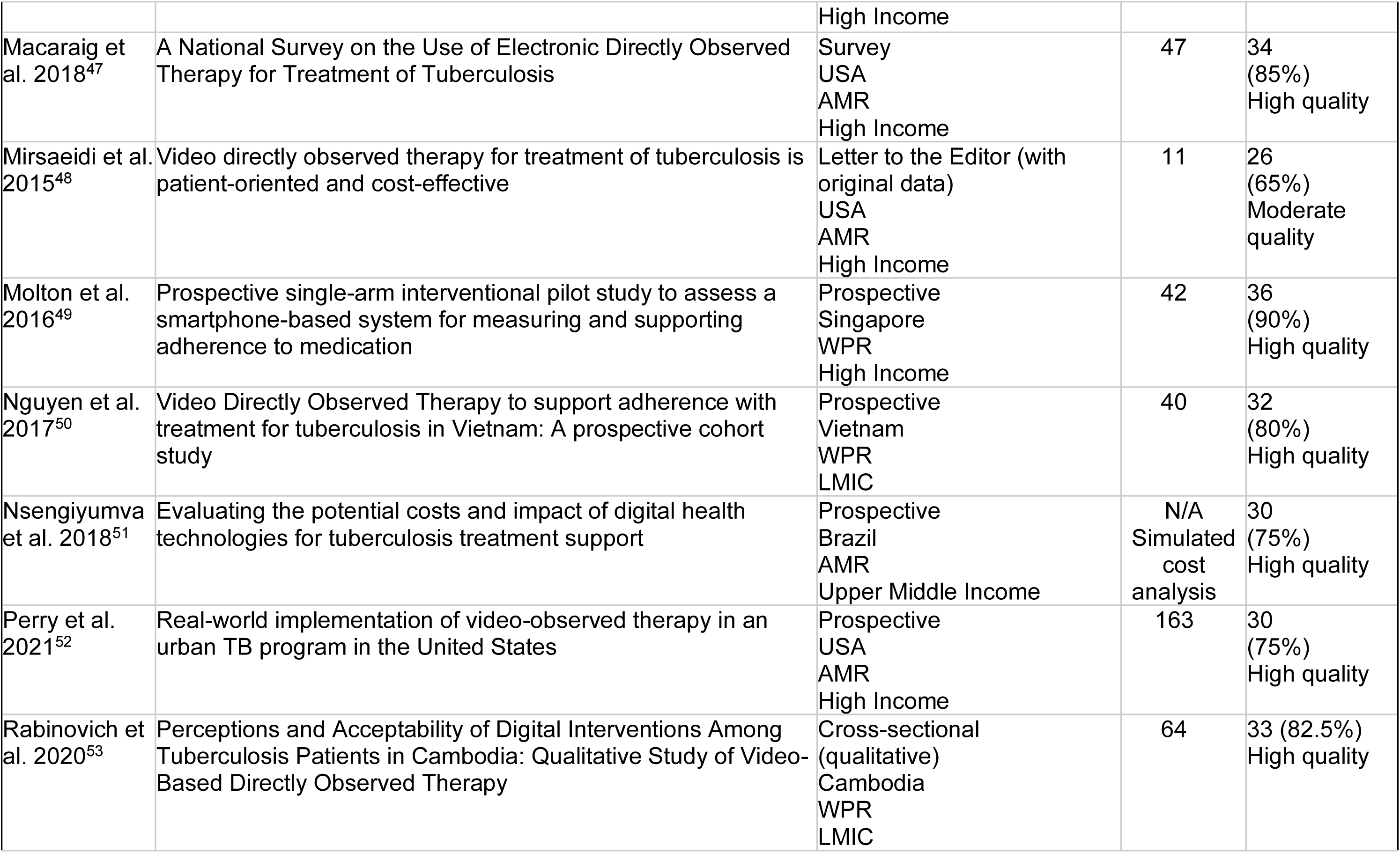
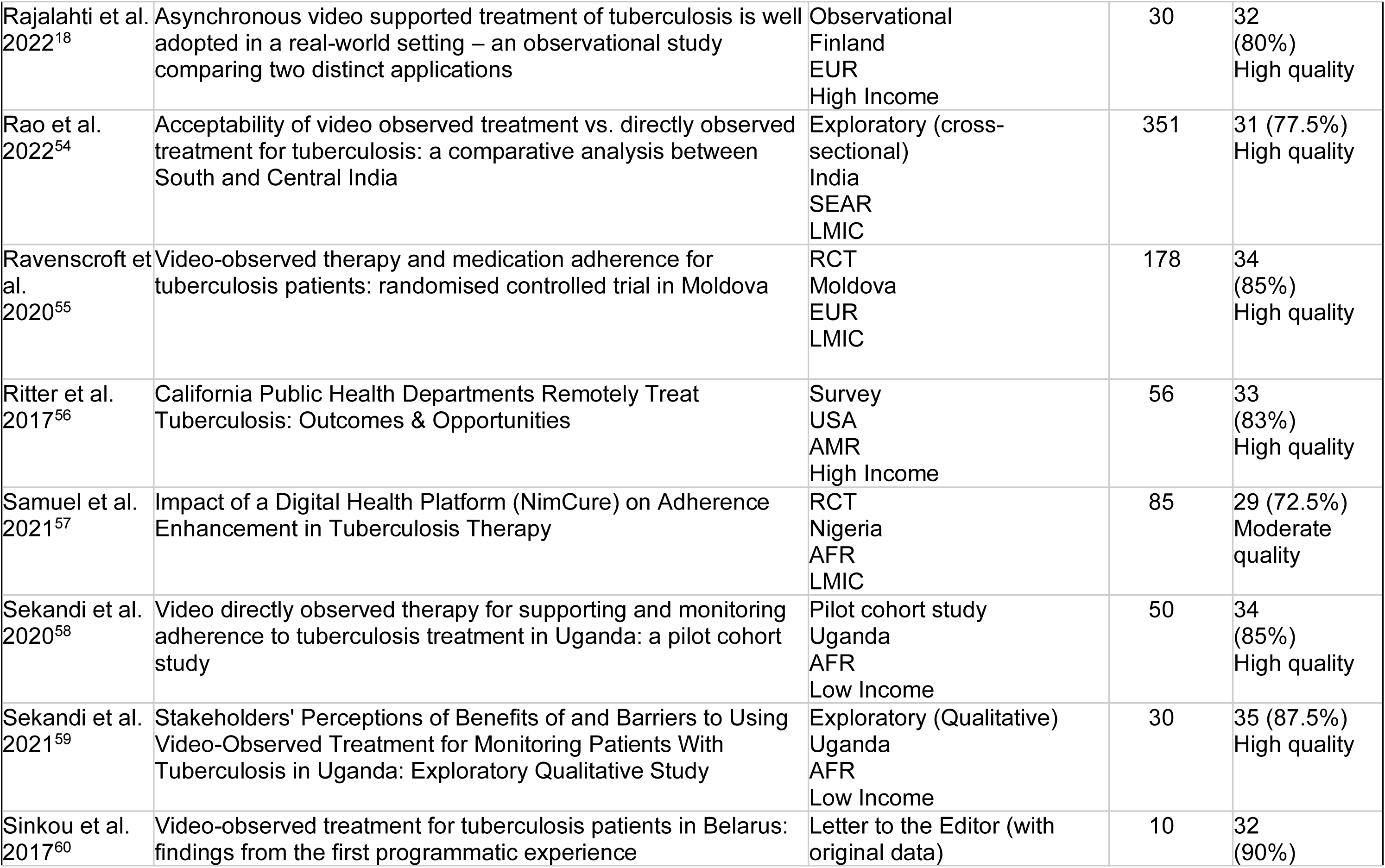
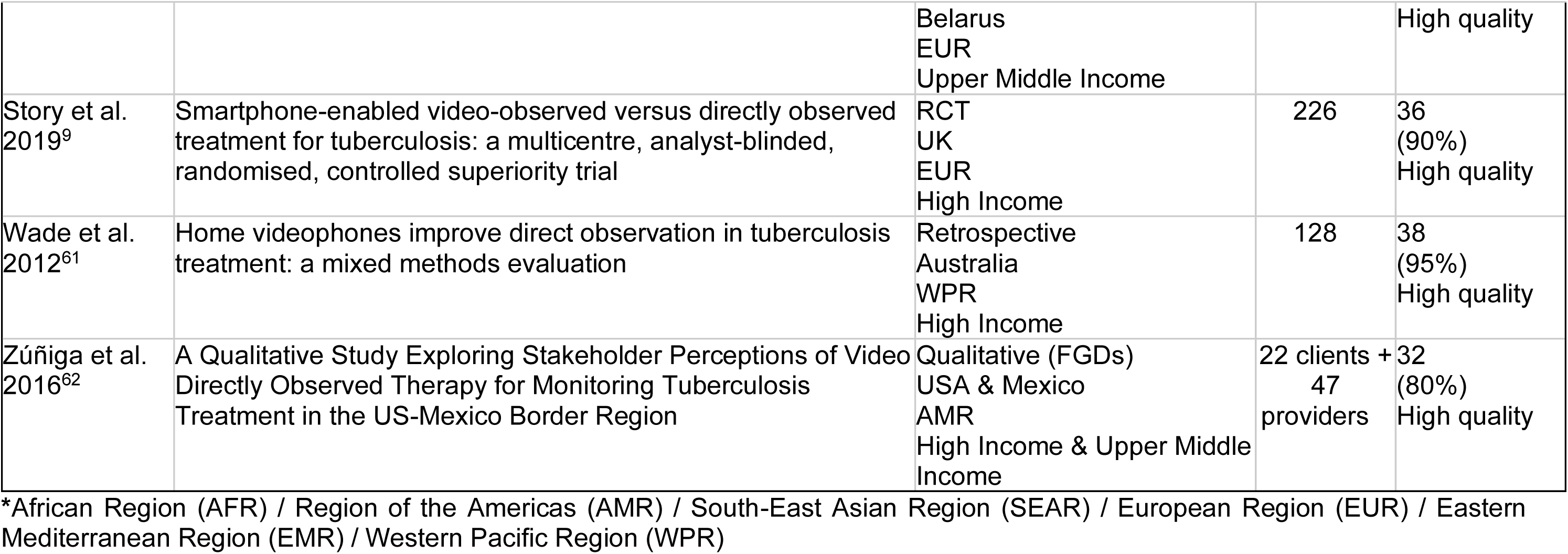
Primary Research Studies (n=47)

**Table 2:** Review-type Studies (n=13)

| Author & Year | Title | Self-reported Study type and Design / Country of 1<sup>st</sup> author / WHO Region* | Studies included (n) | PRISMA or similar reporting guidelines followed | CCAT score (%) and grading |
| --- | --- | --- | --- | --- | --- |
| Abbara et al. 2020<sup>63</sup> | The challenges of tuberculosis control in protracted conflict: The case of Syria | Narrative review<br>UK<br>EUR | Not reported | No | 33 (82.5%)<br>High quality |
| Alipanah et al. 2018<sup>4</sup> | Adherence interventions and outcomes of tuberculosis treatment: A systematic review and meta-analysis of trials and observational studies | Systematic review with Meta-analysis<br>USA<br>AMR | 129 | Yes | 37 (92.5%)<br>High quality |
| Brett et al. 2020<sup>64</sup> | Direct Observational Therapy for the Treatment of Tuberculosis: A Review of Clinical Evidence and Guidelines | Quasi-systematic review<br>Canada<br>AMR | 3x systematic reviews, 6x RCTs, 6x evidence-based guidelines | Yes | 35 (87.5%)<br>High quality |
| Chakaya et al. 2021<sup>65</sup> | Global Tuberculosis Report 2020 – Reflections on the Global TB burden, treatment and prevention efforts | Narrative review<br>Kenya<br>AFR | Not reported | No | 36 (90%)<br>High quality |
| Garfein et al. 2019<sup>5</sup> | Synchronous and asynchronous video observed therapy (VOT) for tuberculosis treatment adherence monitoring and support | Literature review (non-systematic)<br>USA<br>AMR | Not reported | No | 28 (70%)<br>Moderate quality |
| Keutzer et al. 2020<sup>66</sup> | Mobile Health Apps for Improvement of Tuberculosis Treatment: Descriptive Review | Descriptive review<br>Sweden<br>EUR | 55 | Yes | 29 (72.5%)<br>Moderate quality |
| Lee et al. 2020<sup>67</sup> | Use of Digital Technology to Enhance Tuberculosis Control: Scoping Review | Scoping review<br>Switzerland<br>EUR | 145 | Yes | 33 (82.5%)<br>High quality |
| Ngwatu et al. 2018<sup>68</sup> | The impact of digital health technologies on tuberculosis treatment: a systematic review | Systematic review<br>Switzerland<br>EUR | 7 | Yes | 36<br>(90%)<br>High quality |
| Ridho et al. 2022<sup>69</sup> | Digital Health Technologies to Improve Medication Adherence and Treatment Outcomes in Patients With Tuberculosis: Systematic Review of Randomized Controlled Trials | Systematic review of RCTs<br>Indonesia<br>SEAR | 16 | Yes | 37<br>(92.5%)<br>High quality |
| Stoner et al. 2022<sup>8</sup> | Digital directly observed therapy to monitor adherence to medications: a scoping review | Scoping review<br>USA<br>AMR | 28 | Yes | 35<br>(85%)<br>High quality |
| Subbaraman et al. 2018<sup>70</sup> | Digital adherence technologies for the management of tuberculosis therapy: mapping the landscape and research priorities | Narrative review<br>USA<br>AMR | Not reported | No | 33<br>(82.5%)<br>High quality |
| Truong et al. 2022<sup>71</sup> | Video-Observed Therapy Versus Directly Observed Therapy in Patients With Tuberculosis | Systematic review and Meta-analysis<br>USA<br>AMR | 9 | Yes | 35<br>(87.5%)<br>High quality |
| Wong et al. 2022<sup>72</sup> | Digital health use in latent tuberculosis infection care: A systematic review | Systematic review<br>Malaysia<br>WPR | 15 | Yes | 36<br>(90%)<br>High quality |

**Table 3.** : Grey Literature documents (n=6)

| Organisation (author) & Year | Title | Type of document | Country / WHO Region | Current TB burden, cases per 100,000* |
| --- | --- | --- | --- | --- |
| CDC 2017<sup>73</sup> | Implementing an Electronic Directly Observed Therapy (eDOT) Program: A Toolkit for Tuberculosis (TB) Programs | Toolkit | USA AMR | 2.5 |
| ECDC (Sandgren et al.) 2016<sup>74</sup> | Guidance on tuberculosis control in vulnerable and hard-to-reach populations | Guideline | Europe EUR | 10 |
| WHO (Falzon et al.) 2015<sup>10</sup> | Digital health for the end TB strategy: An agenda for action | Strategy | Worldwide | 127 |
| WHO (Farr et al.) 2017<sup>75</sup> | Handbook for the use of digital technologies to support tuberculosis medication adherence | Handbook | Worldwide | 127 |
| WHO 2017<sup>76</sup> | Guidelines for treatment of drug susceptible tuberculosis and patient care | Guideline | Worldwide | 127 |
| WHO Europe 2020<sup>7</sup> | Quick guide to video-supported treatment of tuberculosis | Quick guide | Europe EUR | 10 |

\*Sources: WHO, 2021; ECDC, 2022; CDC, 2023

Most studies (n=34) were conducted at study sites in urban settings, 16 were in suburban/peri-urban and only one study^33^ assessed VOT in the rural context. Of the 55 studies from which a country and income level could be inferred, 33/55 (60%) of the studies were from high-income countries, 8/55 (15%) were from upper-middle-income and middle-income countries, and 14/55 (25%) were from LMIC. Most high-income studies came from the USA (n=25, Supplementary Figure 1), which has a low TB incidence of 2.5 cases per 100,000^77^.

### Intervention Types

Of the 47 primary research studies, the majority (n=29) were single-arm intervention studies of VOT with no comparison group. A minority of studies (n=15) were VOT versus DOT head-to-head comparisons, and the remaining three were VOT compared with other adherence methods such as SMS and medication monitors^51^, comparison of two different VOT apps^18^, and one study examining VOT in a paediatric population^31^. Four studies (4/66, 8%)^9,36,55,57^ were Randomised Control Trials (RCT) and, of the five systematic reviews, two^4,71^ had an accompanying meta-analysis. One study^26^ specifically evaluated VOT for supporting people with MDR-TB. No studies were identified that assessed VOT interventions amongst pregnant women or specifically amongst people with TB/HIV co-infection.

### A Comprehensive Summary of VOT Applications

In the 66 records identified, a wide variety of applications and technologies were mentioned. Of the 20 applications identified, 12/20 (60%), including the top five most cited or used, were developed in the USA. The only VOT application developed in an LMIC context was NIMCURE from Nigeria, which has the highest TB burden in Africa accounting for 4.6% of the global TB cases^78^. Supplementary Table 1 summarises the applications’ characteristics, costs, and which records cited or used these.

### Critical Appraisal using CCAT

Across the primary studies and review-type studies where the CCAT was applicable, the range of scores was from 21 (52.5%) to 39 (97.5%) out of 40, with a mean score of 32.38 (81%). This meant that all studies were rated as moderate to high quality on the CCAT scoring system. See Supplementary Table 2 for the disaggregation of CCAT scores attained.

### Feasibility

A total of 43/66 (65%) records reported on assessments of the feasibility of VOT as a modality to measure TB adherence. Of these, 35 were primary studies, five were review-type studies and three were grey literature documents. All studies reported that VOT was feasible regardless of urbanicity (although most were conducted in urban/peri-urban settings), country income level or population context. However, the most frequently reported challenges to feasibility were lack of or unreliable internet coverage^5,25,26,28,36,39,42,48,50,56,57,61,75^, low quality of videos or incomplete dose verification^17,26,28,34,35,40–42,60,70^ and other technical issues including battery problems^18,22,25,28,31,33,49,50,58^.

Studies identified the need to loan a VOT-compatible smartphone to people with TB^33,34,44,50^ and consider the cost of packages of mobile phone data^5^. One study^53^ found that prior ownership of a smartphone was not associated with the ability to learn how to use VOT. Potential lack of rapport between HCWs and people with TB was described^5,35,62^ and challenges to adherence including not attending scheduled VOT appointments or being lost to follow-up were perceived to be similar to standard DOT^25,26,28,33^.

One retrospective study in Baltimore (urban), USA^46^ reported usage of VOT during the COVID-19 pandemic when DOT was not possible. It highlighted age as the main barrier to the use of VOT, with older participants finding the technology more challenging. Two studies^23,52^ also reported that VOT uptake, was more common in younger people. Fewer than 10% of participants in Guo *et al.*^37^ experienced issues with uploading videos for VOT and Garfein *et al.*^32^ reported that healthcare providers were able to verify most VOT-delivered doses. Holzman *et al.*^40^ showed that VOT was utilised successfully when participants travelled to another country, which the authors interpreted as potentially facilitating increased acceptability of VOT. Ravenscroft *et al*.^55^ highlighted that people with TB using VOT were more likely to report side effects than people with TB using DOT.

VOT was found to be technically feasible in the conflict setting of Syria^63^ but its utility was limited by intermittent supply of TB medications. Donahue *et al.*^31^ found at a military health facility for families of service members found that VOT was feasible in a paediatric population with parental support. A WHO report^10^ called “Digital health for the end TB strategy: An Agenda for action” considered that VOT could become more feasible as coverage of mobile smartphones with internet capability increased.

In its guidance on TB control in Europe, the ECDC scientific panel deemed that VOT was “Likely” feasible to use for vulnerable and hard-to-reach populations^74^.

### Acceptability

A total of 44/66 (66%) records, mostly primary research studies, evaluated and reported on the acceptability of VOT. Except for a single study in the Philippines^26^, in which uptake of VOT was low (36%), all found VOT acceptable to people with TB and healthcare providers. A preference amongst people with TB for VOT over DOT was reported to relate to increased convenience, time-saving, and flexibility^22–25,32–34,36,37,40–42,48,54^. Amongst healthcare workers, two studies from China and The Philippines, reported a preference for VOT over DOT^26,37^. In studies which measured satisfaction levels amongst people with TB using VOT versus people with TB using DOT, people using VOT reported higher levels of satisfaction^27,55,72^.

Both people with TB and healthcare workers or providers raised privacy, security, and confidentiality as potential concerns with VOT across several studies ^5,33,34,36,41,47,48,50,57,58^. Specifically, Garfein *et al.*^5^ identified cultural concerns of videos of females with TB being observed by male care providers in certain countries or contexts. However, this was balanced by studies reporting that both people with TB and HCWs perceived VOT to be more confidential than DOT^17,22,32,34,41,72^. Rabinovich *et al.’s* concept study^53^ stated that although clients were receptive to the prospect of a mobile application for VOT purposes, there were concerns surrounding usability of the technology, especially for people with low literacy or who were illiterate.

In Vietnam, Nguyen *et al.*^50^ noted the most common reason (34.2%) for unwillingness to engage in VOT was not wanting to use a smartphone, but this was not mirrored by other studies. In other studies, people with TB reported appreciated *ad hoc* communication offered by VOT^39,41^ and nurses valued the professional development gained from IT training for VOT.

### Costs

A total of 20/66 (33%) records, including 15 primary studies, examined costs associated with VOT. A further nine studies commented subjectively on the cost-saving attributes of VOT but did not conduct formal costing analyses. Most studies reported that, compared to healthcare facility-based or community-based DOT, VOT was associated with cost savings for people with TB, the health system, or both. There were, however, *two* exceptions: Asay *et al*.^19^ evaluated three TB programs offering VOT and found that, in Rhode Island, USA, a high-income context, VOT costs were not different from community-based DOT. This may have related to power because the same study suggested costs to people with TB and the health system were lower with VOT in larger cities (and cohorts) including in New York City and San Francisco. In a systematic review, Wong *et al.*^72^ found that, in the context of tuberculosis infection, SMS-reminder technology was the most cost-effective method for promoting adherence to TB preventive therapy, quoting a cost-analysis simulation study^51^, based in Brazil, a middle-income, high TB and TB/HIV country context.

Cost-saving attributes of VOT over DOT were due to reduced travel costs and reduced time spent observing doses in-person, with studies commenting specifically on personnel (staff) costs^5,9,17,24,33,43,48,61^, hours and costs including transport saved by people with TB^5,37,48,55^. Chuck *et al.*^28^ found that staff could observe twice the number of people with TB taking their medications per allotted appointment slot duration when using VOT compared with DOT. Asynchronous VOT was found by Lam *et al*^45^ to have lower costs to the health system than synchronous VOT but a mobile phone, software purchase, installation costs, and costs to people were not evaluated.

### Effectiveness

A total of 39/66 (59%) records commented on VOT effectiveness, 29 of which were primary research studies, nine were review-type studies, and one was a grey literature article. Versus DOT, VOT was reported to be associated with better adherence across various settings^8,9,21,27,28,30,33,37,46,52,55,68,71^. This was often interpreted as relating to VOT, especially asynchronous VOT, being provided 7-days per week and being less affected by staffing shortages, strikes, and national holidays. Several studies reported no difference in adherence rates between VOT and DOT^5,17,22,35,40,47,57,64^. Perry *et al.*^52^ reported more missing doses (average 3.4% vs. 1.3%, respectively; P < 0.001) over a median period of 27 weeks but similar adherence rates (96% vs. 90%; P=0.326) in VOT vs DOT. Several studies reported high adherence rates with VOT but without comparison to a DOT control group^18,24,31,32,34,41,48,50,58^.

Another common measure of effectiveness was treatment completion as defined by the WHO^76^, which was reported as higher with VOT than DOT in various studies^8,27,44^. Comparable treatment completion rates with VOT versus DOT were reported in other studies^4,26,28,36,47,68,71,76^. No study reported treatment completion rates to be worse amongst people using VOT compared to DOT but, notably, none were specifically designed to demonstrate treatment completion inferiority or superiority. Mortality was infrequently reported but was found to be comparable in three studies^4,36,76^ and, in the study by Casalme *et al.*^26^ amongst people with MDR-TB, mortality was found to be lower with VOT compared with DOT. In some studies, loss to follow-up was either lower^26^ or the same^36^ with VOT versus DOT. No evidence of effectiveness to reduce other adverse TB treatment outcomes including treatment failure were identified.

Treatment success was higher amongst people with TB using VOT than DOT in one study^26^, but comparable between VOT and DOT in other studies^36,52,55,64^. Culture conversion rates for VOT were similar to DOT in three studies^37,52,71^. TB recurrence (i.e. return of symptoms and signs including radiological signs of clinical TB and/or microbiologically-confirmed TB) was reported in Doltu *et al.*^30^ and Guo *et al.*^37^ in the DOT control groups but not VOT groups. No other studies reported TB recurrence in either the VOT or DOT groups.

The key success, challenges, and potential solutions to VOT implementation identified above are summarised and disaggregated by country income in Table 4.

**Table 4:** Reported Successes and Challenges in High-income to Middle-income Countries.

| High-income to Middle-income |  |  |  |
| --- | --- | --- | --- |
| Successes | Challenges | Potential solutions | Key studies |
| Reported high adherence rates with VOT<br>VOT was feasible for people living with TB during the COVID-19 pandemic<br>VOT was feasible for people with (M)DR-TB<br>VOT was feasible for TB preventive therapy monitoring in a paediatric population<br>VOT uptake in individuals older than 65 was 10% in an urban setting in the USA<br>Ability to learn VOT was not associated with ownership of a smartphone<br>When scaled-up in urban settings of the USA, VOT was cost saving compared with DOT<br>HCWs preferred VOT over community DOT home visits<br>VOT was implemented more widely in London, UK based on outcomes from the trial conducted by Story et al. 2019 | Health insurance policies and frameworks must recognise VOT as a treatment in order that patients have their costs reimbursed<br>Poor video quality<br>Network connectivity problems<br>Difficulty navigating VOT software / uploading videos<br>Personal smartphone not compatible for VOT software<br>Lack of interpersonal interaction reported by both clients and HCW<br>Misuse of loaned phone for non-VOT activities<br>Side-effects may go unreported with VOT usage<br>Concerns regarding privacy and data security<br>Older people, males, people with fewer years of formal education, and people with lower annual income were less likely to own smartphones posing problems for equity of access and use | Advocate for recognition of VOT as an established adherence monitoring modality, including both to National TB Programs but also to social protection and health insurance schemes and policy makers<br>Training of HCW and clients<br>Loaning smartphones compatible with VOT software<br>Encourage ad hoc communication and reporting of side-effects via VOT and/or integration of adverse effect monitoring or interactive platforms within VOT applications<br>Reassure clients regarding privacy and data security | Gassanov, 2013<sup>35</sup><br>Chuck, 2016<sup>28</sup><br>Ritter, 2017<sup>56</sup><br>Garfein, 2018<sup>33</sup><br>Lam, 2018<sup>44</sup><br>Macaraig, 2018<sup>47</sup><br>Story, 2019<sup>9</sup><br>Asay, 2020<sup>20</sup><br>Bendiksen, 2020<sup>22</sup><br>Bommakanti, 2020<sup>23</sup><br>Garfein, 2020<sup>34</sup><br>Donahue, 2021<sup>31</sup><br>Perry, 2021<sup>52</sup><br>Bachina, 2022<sup>21</sup><br>Burzynski, 2022<sup>25</sup> |
| Low-income to lower-middle income |  |  |  |
| Successes | Challenges | Potential solutions | Key studies |
| VOT was feasible in LMIC and LIC contexts | Personal preference for DOT<br>Persistent non-adherence with VOT requiring switch back to DOT | Improve the evidence base on implementation strategies, scalability, effectiveness (vs DOT for adherence | Hoffman, 2010<sup>39</sup><br>Nguyen, 2017<sup>50</sup> |
| • VOT was feasible for people living with MDR-TB in an LMIC context (The Philippines)<br>• Positive feedback from clients and HCWs for VOT<br>• Clients could feedback treatment-related problems out of hours using asynchronous VOT<br>• Asynchronous VOT had no sessional time limits<br>• Clients felt empowered by ad hoc communication offered by asynchronous VOT<br>• Opportunity for HCWs to engage in Continued Professional Development in learning requisite IT skills for VOT | • Client unease with recording themselves taking their medications<br>• Concerns regarding privacy following uploading of video-recording<br>• Lack of private place to record VOT videos<br>• Perceived lack of contact with HCW expressed in FGD about VOT in Cambodia<br>• Slow uploading of asynchronous VOT videos due to issues with<br>• Poor network connection, especially in rural areas<br>• Competing for internet bandwidth with the host hospital to deliver VOT<br>• Study smartphone not returned despite expectations it would be so at end of study implementation<br>• Broken or uncharged smartphone | and treatment success), and cost effectiveness of VOT and disseminate to clients, potential clients, public, and HCWs<br>• Reassure clients regarding privacy and data security, consider encryption similar to other platforms including pins, signatures, and digital prints to access<br>• Encourage ad hoc communication and reporting of side-effects via VOT and/or integration of adverse effect monitoring or interactive platforms within VOT applications<br>• Help transition VOT clients between TB centres (if feasible)<br>• Lobby stakeholders for wider mobile internet network reach into rural communities | Holzman, 2019<sup>41</sup><br>Rabinovich, 2020<sup>53</sup><br>Sekandi, 2020<sup>58</sup><br>Samuel, 2021<sup>57</sup><br>Sekandi, 2021<sup>59</sup><br>Casalme, 2022<sup>26</sup><br>Rao, 2022<sup>54</sup> |

## Discussion

The findings from this scoping review illustrate a lack of evidence, especially from randomised controlled trials, on the feasibility, acceptability, cost and effectiveness of VOT for people with TB in LMIC. In predominantly HIC, VOT was shown to contribute to high adherence levels and to have the potential to be more effective and cost-saving than DOT. A wide variety of VOT applications were used, most of which were developed in HICs. International consensus guidelines relating to VOT from WHO, ECDC, and CDC, were largely based on best practice and with low-grade evidence.

### Availability of VOT

This scoping review was able to build on the findings of a scoping review from 2016^79^ to update and summarise the current landscape and diversity of VOT applications used and their pricing, availability, and user interface/platform. It was identified that costs associated with VOT applications, such as user licenses, downloads, and recurrent subscription costs, are not transparent or easily available. In addition, some study trials provided smartphones for participants which may not be sustainable or suitable for upscaling within the health systems of many LMIC high TB burden settings beyond trial conditions.

### Feasibility

This scoping review found that VOT was uniformly reported to be *feasible* in HIC but more evidence is needed from LMIC. VOT is now an established alternative to DOT in New York City, USA and London, UK^9,28^. Shared challenges to feasibility were identified that commonly related to technical aspects such as internet access, submissions of poor-quality videos, and initial problems with navigating the VOT applications, but most studies were able to overcome these without significant loss to follow-up reported. Personal access to a smartphone or loan of a smartphone to participants by the study was an accessibility issue raised^11,23^. For TB programmes to sustain VOT delivery, a pre-intervention baseline analysis of the use of smartphones and network coverage and consideration of underserved groups, including those without access, extremely impoverished, or with literacy and digital literacy issues, is essential to ensure equitable delivery of VOT, echoing recommendations from Peru^80^ and WHO^75^.

### Acceptability

People living with TB and healthcare workers were widely receptive to VOT as an acceptable alternative to DOT. The WHO “Handbook for the use of digital technologies to support tuberculosis medication adherence”^75^ suggested the need to evaluate acceptability of VOT in subpopulations such as adolescent girls and women in high TB burden LMIC to ensure equitable impact. The ECDC scientific panel rated VOT as “likely acceptable” for adherence monitoring of underserved populations in Europe^74^. Future VOT implementations must assure participants that their privacy will be maintained and ensure data security in line with local legislative requirements, such as the UK’s Data Protection Act 2018. In addition, differentiating the process and impact outcomes of asynchronous versus synchronous VOT remains a knowledge gap.

To enhance acceptability for HCWs, VOT training should be used as an opportunity for continued professional development of IT skills. From the perspective of people with TB, it must be recognised that VOT is not a blanket solution for all and TB programmes should retain the option for DOT for those who prefer or require face-to-face interactions.

### Costs

Of the minority of studies that formally collected and analysed cost data, most reported cost savings of VOT versus DOT for people with TB and the health system. The actual and proportional cost savings may vary between HIC and LMIC settings due to differential personnel, equipment, and logistics costs. For future studies, it was noted that other costs such as incentivising adherence, for example through weekly food vouchers, must also be included for accurate cost calculations. Our findings were similar to a previous scoping review^67^, which also found cost-savings of up to 58% for the healthcare system amongst 19 studies and in agreement with the ECDC scientific panel contribution^74^ that VOT was “Likely” cost-effective.

For people with TB, the most widely reported cost saving of VOT avoiding travel costs to DOT clinics, was also noted to have potential positive public health implications^81^. For HCWs, VOT was reported to improve time-efficiency and, in some cases, reduce their own travel costs compared to DOT.

Some important gaps in the literature relating to cost of VOT include within trial cost-effectiveness analyses, differential costs between *synchronous* and *asynchronous* VOT, and incorporating the impact of ownership, loans, or access to internet-enabled smartphone devices.

### Effectiveness

The overarching evidence from diverse studies including early trials and post-implementation evaluations suggests that treatment outcomes in VOT are likely to be at least non-inferior to DOT. Other outcomes including treatment completion, bacteriological resolution, and mortality rate were similar to standard DOT. In line with these findings, WHO previously concluded that VOT was at least non-inferior to DOT in terms of TB treatment completion and mortality^76^. However, it is clear that large-scale, randomised-controlled trials in diverse settings, especially in LMIC or with vulnerable populations including those with MDR-TB, are required.

### Strengths & Limitations

This scoping review has updated, consolidated and synthesised the existing evidence base on the use of VOT – including a novel table summarising the VOT applications available - to support people with TB globally. The methods are repeatable and the quality scoring using CCAT is transparent.

The review had several limitations. First, reviewers were not blinded, risking bias, and the search criteria did not include studies prior to 1st January 2000. However, the subsequent snowballing of citations did not find relevant studies earlier than this date. We also found multiple international clinical trial protocols for VOT evaluation on trials registration sites that have yet to publish their results. Second, several studies acknowledged selection bias in their recruitment with few involving randomisation, which limits their generalizability and warrants caution in the interpretation of their findings. Third, there was limited evidence evaluating forms of VOT such as synchronous versus asynchronous, the platforms or applications used to implement VOT, and vulnerable, underserved groups were neglected with only one study of VOT in children and none focused on pregnant women or people with TB and HIV or other comorbidities. These are notable gaps in the literature and understanding how VOT could be adapted for these populations is crucial for a comprehensive TB programme service delivery.

### Research Recommendations

There is significant potential for future research to move the field of VOT for TB forward, especially in high TB burden LMIC. Randomised-controlled trials including those with multi-arm multi-stage or hybrid effectiveness-implementation designs and incorporating distributional or extended cost-effectiveness analyses and recognised WHO TB Patient Cost Survey methods^77^ could be of great value to evaluate not only effectiveness but also feasibility, acceptability, cost-effectiveness and equity of VOT. However, RCTs can be expensive and time-consuming and, programmatically, other studies or implementation projects including the use of quasi-experimental designs or interrupted time-series analysis may be more suitable.

Wider research including ecological studies and secondary analysis of existing household survey data, including data on access to phones and internet, could examine how increased penetrance of smartphone coverage and usage in LMIC could contribute to VOT being integrated within standard of care. Moreover, there has been minimal exploration of sociocultural factors associated with mobile phone usage and ownership, particularly in LMIC where there may be a gender differential in household-level economic agency. Understanding such sociocultural factors can impact VOT feasibility and hence be crucial for optimising its effectiveness. Future studies implementing and evaluating VOT would benefit from considerations of equity within their analyses, including disaggregating data by gender, urban versus rural location, socioeconomic position, educational level, literacy, and digital literacy.

Finally, broader potentials of internet-enabled smartphone technology in the future could be, alongside VOT, screening for TB in households, for example via acoustic cough analysis,^82^ monitoring TB preventive therapy adherence and completion, reviewing symptoms of chronic respiratory diseases in people with suspected TB but no formal diagnosis, and adjunctive features to monitor side-effects over the course of a treatment regimen.

## Conclusion

The COVID-19 pandemic has accelerated acceptance of digital health technologies, their implementation, and utility. This scoping review found that VOT was reported to be a feasible, acceptable, and effective alternative to DOT, which was likely cost-saving to people with TB and health systems in HIC. More evidence is needed to inform its implementation and potential scale-up in LMICs with high TB burden.

## Data Availability

The data are available from the corresponding author (email address) upon reasonable request.

## Contributions

An Du Thinh (ADT): Conceptualization, Methodology, Investigation, Data Curation, Formal analysis, Writing - Original Draft, Review & Editing, Visualization

Eleanor Morgan (EM): Conceptualization, Methodology, Investigation, Data Curation, Formal analysis, Writing – supported Original Daft, Review & Editing, Visualization Kassia Pereira (KP): Conceptualization, Methodology, Investigation, Data Curation, Formal analysis, Writing - Review & Editing, Visualization

Celso Khosa (CK): Conceptualization, Methodology, Investigation, Formal analysis, Writing - Review & Editing, Visualization

Tom Wingfield (TW): Conceptualization, Methodology, Investigation, Formal analysis, Writing - Review & Editing, Visualization

All authors read and approved the final manuscript.

## Ethical Considerations

There were no ethical issues or risk of harm to patients identified. Ethical approval exemption was sought and granted by LSTM. All the resources used were based on published research with previously anonymised participant demographics and outcomes. This study was nested within the larger “SAFEST-1 MDR-TB” programme of research, which includes an implementation study of VOT for people with MDR-TB in Mozambique. SAFEST-1 MDR-TB was approved by Research Ethics Committees at the Liverpool School of Tropical Medicine, UK (23-008) and the Comité Institucional de Ética do Instituto Nacional de Saude, Mozambique (148/CIE-INS/2023, 15^th^ of November, 2023).

## Availability of data and materials

The data are available from the corresponding author (email address) upon reasonable request.

## Appendix/Supplemental Materials

An example of the MeSH (Medical Subject Headings) search terms used were: (TI (VOT OR (video AND observed AND therapy) OR VST OR (video AND support AND treatment) OR vDOT OR eDOT)) AND (TI (TB OR tuberculosis)) OR (AB (VOT OR (video AND observed AND therapy) OR VST OR (video AND support AND treatment) OR vDOT OR eDOT)) AND (AB (TB OR tuberculosis)). These MeSH terms were piloted and refined between team members ADT, EM, and KP.

### Abbreviations and Glossary

ADT: An Du Thinh
AMR: Antimicrobial resistance
CCAT: Crowe Critical Appraisal Tool
CDC: Centers for Disease Control and Prevention; American public health agency
CI: Confidence Interval
CK: Celso Khosa
Client: Person living with TB; a less stigmatising term than “TB patient”
CXR: Chest x-ray
DOT: Direct Observed Therapy; commonly associated with in-person adherence monitoring for TB treatment
DS-TB: drug-sensitive TB
ECDC: European Centre for Disease Prevention and Control - the European public health agency
eDOT: electronic-Direct Observed Therapy; VOT in a different name
EM: Eleanor Morgan
FGD: Focus Group Discussions – structured group interviews to collect qualitative data
GRADE: Grading of Recommendations, Assessment, Development, and Evaluations framework for evaluating research evidence summaries
HCW: Healthcare Worker
IGRA: Interferon-gamma release assay
INS: Instituto Nacional de Saúde
JBI: Joanna Briggs Institute; consensus framework for conducting Scoping Reviews
KP: Kassia Pereira
LMIC: Lower Middle Income Country as determined by The World Bank classification
LSTM: Liverpool School of Tropical Medicine
LTBI: Latent Tuberculosis Infection
MDR-TB: Multidrug Resistant Tuberculosis
MTB: Mycobacterium tuberculosis
NHS: National Health Service
NTP: National Tuberculosis Programme
PRISMA: Preferred Reporting Items for Systematic Reviews and Meta-Analyses
RCT: Randomised Control Trial
RR: Relative Risk
SAT: Self-Administered Therapy; TB treatments taken unobserved or monitored by healthcare professionals
ScR: Scoping Review; as in PRISMA-ScR
SDG: Sustainable Development Goals as set by the WHO/UN
SMS: Short Message/Messaging Service
TB: Tuberculosis
TST: Tuberculin skin test
TW: Tom Wingfield
vDOT: virtual/video-Directed Observed Therapy; VOT in a different name
VOT: Video Observed Therapy; also known as Video-Supported Treatment, electronic-Directly Observed Therapy and virtual/video-Directly Observed Therapy
VST: Video-Supported Treatment; VOT in a different name
WHO: World Health Organisation
UKHSA: United Kingdom Health Security Agency; agency formerly known as Public Health England (PHE)

---

**Supplementary Table 1:** Summary of VOT Applications.

| Name | Website & App stores | Country of Origin / WHO Region | Cost | Cited or used by |
| --- | --- | --- | --- | --- |
| Emocha Health, now renamed Scene Health | www.scene.health or via Apple App Store and Google Play Store | USA<br>AMR | Free to download the app, but the cost of the account subscription cost to on Scene Health not reported | Bachina, 2022<sup>21</sup>; Perry, 2021<sup>52</sup>; Lippincott, 2022<sup>46</sup>; Holzman, 2019<sup>41</sup>; Holzman 2018<sup>40</sup>; Keutzer 2020<sup>66</sup>; Asay, 2020<sup>20</sup>; CDC, 2017<sup>73</sup>; Farr, 2017<sup>75</sup> |
| SureAdhere | www.sureadhere.com owned by Dimagi; Apple App Store and Google Play Store | USA<br>AMR | Starting from \$120/month | Garfein, 2019<sup>5</sup>; Casalme 2022<sup>26</sup>; Keutzer, 2020<sup>66</sup>; Asay, 2020<sup>20</sup>; Nguyen, 2017<sup>50</sup>; CDC, 2017<sup>73</sup>; Farr, 2017<sup>75</sup>; Subbaraman, 2018<sup>70</sup> |
| Skype | www.skype.com owned by Microsoft; Apple App Store and Google Play Store | USA<br>AMR | Free for internet calls only, variable costs otherwise dependent on the country | Asay, 2020<sup>20</sup>; Mirsaeidi, 2015<sup>48</sup>; Macaraig, 2018<sup>47</sup>; CDC, 2017<sup>73</sup>; Buchman, 2017<sup>24</sup> |
| FaceTime | Pre-installed on Apple mobile devices; Apple App Store only, owned by Apple | USA<br>AMR | Free for calls between Apple devices | Macaraig, 2018<sup>47</sup>; CDC, 2017<sup>73</sup> |
| AiCure | www.aicure.com Apple App store and Google Play store | USA<br>AMR | Starting from \$750 per clinic per month for up to 50 patients (Salcedo et al., 2021) | Keutzer, 2020<sup>66</sup>; CDC, 2017<sup>73</sup> |
| Acano | Via Apple App Store only, owned by Cisco Systems | USA<br>AMR | Free | Bendiksen, 2020<sup>22</sup> |
| Fuze | www.fuze.com owned by 8x8 Company; Apple App Store and Google Play Store | USA<br>AMR | Starting from \$15 per user per month | CDC, 2017<sup>73</sup> |
| HipaaBridge | www.everbridge.com and Google Play Store | USA<br>AMR | Free to download on Android, unclear from Ever Bridge website if there are other costs from website applicable following download | CDC, 2017<sup>73</sup> |
| ooVoo | www.oovoo.com but discontinued November 2017 | USA<br>AMR | Not reported | CDC, 2017<sup>73</sup> |
| Tango | www.tango.me Apple App Store and Google Play Store | USA<br>AMR | Free to download, in-app purchases for extra features | CDC, 2017<sup>73</sup> |
| Adobe Connect | www.adobe.com or via Apple App Store and Google Play Store | USA<br>AMR | Free | Donahue, 2021<sup>31</sup> |
| ViaTV units<br>*earliest example of VOT amongst included studies | Not commercially available; developed by the research team to prove earliest VOT concept | USA<br>AMR | Not reported | DeMaio, 2001<sup>17</sup> |
| Youping Technology Co. | Not commercially available, developed for study trial | China<br>WPR | Not reported | Guo, 2020<sup>36</sup> |
| MIST (Mobile Interactive Supervised Therapy) | Not commercially available, developed for study trial | Singapore<br>WPR | Not reported | Molton, 2016<sup>49</sup> |
| I Like VST by TBVOT.MD | www.tbvot.md and Google Play Store | Moldova<br>EUR | Free | WHO, 2020<sup>7</sup> |
| Lekar | Via Apple App Store and Google Play, owned by ABM Cloud | Ukraine<br>EUR | Free | WHO, 2020<sup>7</sup> |
| HealthFox | www.healthfox.fi or via Apple App Store and Google Play Store | Finland<br>EUR | Free | Rajalahti, 2022<sup>18</sup> |
| AppVST | Not commercially available, developed by TUH technology department | Finland<br>EUR | Not reported | Rajalahti, 2022<sup>18</sup> |
| NIMCURE | Not commercially available; developed by Co-Creation Hub and Nigerian Institute of Medical Research (NIMR) | Nigeria<br>AFR | Not reported | Samuel, 2021<sup>57</sup> |
| Adhere 2TX-TB | Not reported | Not reported | Not reported | WHO, 2020<sup>7</sup> |

\*Applications developed or evaluated prior to the year 2000 that did not meet the study eligibility criteria for inclusion may exist. All websites accessed January 2024.

**Supplementary Table 2:** Crow Critical Appraisal Tool (CCAT) Breakdown of Scores - Primary & Review Studies (n = 60) Scoring rubric: 0 – 49% = low quality; 50-74% = moderate quality; 75-100% = high quality Adapted from Nuttall et al. 2022^6^

| Study ref. | Preliminaries | Intro | Design | Sampling | Data collection | Ethical matters | Results | Discussion | Total CCAT Score (n/40)<br>/ Grading |
| --- | --- | --- | --- | --- | --- | --- | --- | --- | --- |
| Abbara et al. 2020<sup>63</sup> | 4 | 5 | 4 | 4 | 3 | 5 | 4 | 4 | 33 (82.5%) High Quality |
| Aisyah et al. 2020<sup>19</sup> | 4 | 5 | 4 | 3 | 4 | 5 | 4 | 4 | 33 (82.5%) High Quality |
| Alipanah et al. 2018<sup>4</sup> | 4 | 4 | 5 | 5 | 5 | 5 | 5 | 4 | 37 (92.5%) High Quality |
| Asay et al. 2020<sup>11</sup> | 4 | 5 | 4 | 4 | 3 | 4 | 4 | 5 | 33 (82.5%) High Quality |
| Bachina et al. 2022<sup>21</sup> | 4 | 5 | 4 | 4 | 3 | 4 | 4 | 4 | 32 (80%) High Quality |
| Bendiksen et al. 2020<sup>22</sup> | 4 | 4 | 4 | 3 | 4 | 3 | 4 | 4 | 30 (75%) High Quality |
| Bommakan ti et al. 2020<sup>23</sup> | 4 | 4 | 4 | 3 | 5 | 5 | 4 | 4 | 33 (82.5%) High Quality |
| Brett et al. 2020<sup>64</sup> | 4 | 5 | 5 | 4 | 3 | 4 | 5 | 5 | 35 (87.5%) High Quality |
| Buchman et al. 2017<sup>24</sup> | 4 | 5 | 3 | 5 | 3 | 2 | 3 | 4 | 29 (73%) Moderate Quality |
| Burzynski et al. 2022<sup>25</sup> | 4 | 5 | 5 | 5 | 3 | 5 | 4 | 5 | 36 (90%) High Quality |
| Casalme et al. 2022<sup>26</sup> | 4 | 4 | 4 | 3 | 1 | 5 | 4 | 4 | 29 (72.5%) Moderate Quality |
| Chakaya et al. 2021<sup>65</sup> | 4 | 4 | 4 | 5 | 5 | 5 | 4 | 5 | 36 (90%) High Quality |
| Chen et al. 2020<sup>27</sup> | 4 | 4 | 4 | 3 | 3 | 5 | 4 | 4 | 31 (77.5%) High Quality |
| Chuck et al. 2016<sup>28</sup> | 4 | 5 | 4 | 3 | 3 | 4 | 4 | 4 | 31 (77.5%) High Quality |
| DeMaio et al. 2001<sup>17</sup> | 4 | 4 | 4 | 3 | 3 | 3 | 4 | 4 | 29 (72.5%) Moderate Quality |
| Do et al. 2019<sup>29</sup> | 4 | 5 | 3 | 4 | 4 | 5 | 4 | 4 | 33 (82.5%) High Quality |
| Doltu et al. 2021<sup>30</sup> | 4 | 5 | 4 | 3 | 4 | 5 | 4 | 4 | 33 (82.5%) High Quality |
| Donahue et al. 2021<sup>31</sup> | 4 | 4 | 4 | 3 | 4 | 3 | 4 | 4 | 30 (75%) High Quality |
| Gassanov et al. 2013<sup>35</sup> | 3 | 3 | 3 | 3 | 3 | 0 | 3 | 3 | 21 (52.5%) Moderate Quality |
| Garfein et al. 2015<sup>32</sup> | 4 | 4 | 5 | 4 | 4 | 5 | 4 | 5 | 35 (87.5%) High Quality |
| Garfein et al. 2018<sup>33</sup> | 4 | 4 | 5 | 3 | 5 | 4 | 4 | 4 | 33 (82.5%) High Quality |
| Garfein et al. 2019<sup>5</sup> | 4 | 4 | 3 | 0 | 4 | 5 | 4 | 4 | 28 (70%) Moderate Quality |
| Garfein et al. 2020<sup>34</sup> | 4 | 5 | 4 | 3 | 3 | 5 | 3 | 4 | 31 (77.5%) High Quality |
| Guo et al. 2020a<sup>36</sup> | 4 | 4 | 4 | 3 | 4 | 5 | 4 | 4 | 32 (80%) High Quality |
| Guo et al. 2020b<sup>37</sup> | 4 | 4 | 3 | 3 | 4 | 3 | 4 | 4 | 29 (72.5%) Moderate Quality |
| Hira et al. 2022<sup>38</sup> | 4 | 5 | 4 | 4 | 4 | 0 | 4 | 3 | 28 (70%) Moderate Quality |
| Hoffman et al. 2010<sup>39</sup> | 4 | 5 | 4 | 3 | 3 | 4 | 3 | 3 | 29 (72.5%) Moderate Quality |
| Holzman et al. 2018<sup>40</sup> | 4 | 5 | 4 | 5 | 3 | 5 | 4 | 4 | 34 (85%) High Quality |
| Holzman et al. 2019<sup>41</sup> | 4 | 5 | 4 | 4 | 3 | 4 | 5 | 5 | 34 (85%) High Quality |
| Keutzer et al. 2020<sup>66</sup> | 4 | 4 | 3 | 4 | 3 | 3 | 4 | 4 | 29 (72.5%) Moderate Quality |
| Krueger et al. 2010<sup>43</sup> | 4 | 5 | 4 | 3 | 3 | 3 | 3 | 3 | 28 (70%) Moderate Quality |
| Kumar et al. 2019<sup>42</sup> | 4 | 5 | 4 | 4 | 4 | 5 | 4 | 4 | 34 (85%) High Quality |
| Lam et al. 2018<sup>44</sup> | 4 | 5 | 4 | 3 | 5 | 4 | 4 | 4 | 33 (82.5%) High Quality |
| Lam et al. 2019<sup>45</sup> | 4 | 5 | 4 | 3 | 3 | 5 | 4 | 4 | 32 (80%) High Quality |
| Lee et al. 2020<sup>67</sup> | 4 | 5 | 4 | 5 | 3 | 3 | 4 | 5 | 33 (82.5%) High Quality |
| Lippincott et al. 2022<sup>46</sup> | 4 | 5 | 4 | 4 | 3 | 5 | 4 | 5 | 34 (85%) High Quality |
| Macaraig et al. 2018<sup>47</sup> | 4 | 5 | 4 | 5 | 4 | 4 | 4 | 4 | 34 (85%) High Quality |
| Mirsaeidi et al. 2015<sup>48</sup> | 3 | 4 | 3 | 3 | 3 | 4 | 3 | 3 | 26 (65%) Moderate Quality |
| Molton et al. 2016<sup>49</sup> | 4 | 5 | 4 | 5 | 4 | 5 | 4 | 5 | 36 (90%) High Quality |
| Nguyen et al. 2017<sup>50</sup> | 4 | 5 | 4 | 5 | 3 | 4 | 4 | 3 | 32 (80%) High Quality |
| Ngwatu et al. 2018<sup>68</sup> | 4 | 5 | 5 | 5 | 5 | 4 | 4 | 4 | 36 (90%) High Quality |
| Nsengiyumva et al. 2018<sup>51</sup> | 4 | 4 | 4 | 3 | 3 | 4 | 4 | 4 | 30 (75%) High Quality |
| Perry et al. 2021<sup>52</sup> | 4 | 5 | 3 | 3 | 3 | 4 | 4 | 4 | 30 (75%) High Quality |
| Rabinovich et al. 2020<sup>53</sup> | 4 | 5 | 4 | 3 | 5 | 4 | 4 | 4 | 33 (82.5%) High Quality |
| Rajalahti et al. 2022<sup>18</sup> | 4 | 4 | 4 | 3 | 4 | 5 | 4 | 4 | 32 (80%) High Quality |
| Rao et al. 2022<sup>54</sup> | 4 | 5 | 4 | 3 | 3 | 4 | 4 | 4 | 31 (77.5%) High Quality |
| Ravenscroft et al. 2020<sup>55</sup> | 4 | 5 | 4 | 5 | 3 | 5 | 4 | 4 | 34 (85%) High Quality |
| Ridho et al. 2022<sup>69</sup> | 5 | 4 | 4 | 5 | 5 | 4 | 5 | 5 | 37 (92.5%) High Quality |
| Ritter et al. 2017<sup>56</sup> | 3 | 5 | 4 | 5 | 5 | 2 | 4 | 5 | 33 (83%) High Quality |
| Samuel et al. 2021<sup>57</sup> | 4 | 4 | 4 | 3 | 3 | 4 | 4 | 3 | 29 (72.5%) Moderate Quality |
| Sekandi et al. 2020<sup>58</sup> | 4 | 5 | 4 | 3 | 4 | 5 | 4 | 5 | 34 (85%) High Quality |
| Sekandi et al. 2021<sup>59</sup> | 4 | 5 | 4 | 3 | 5 | 5 | 4 | 5 | 35 (87.5%) High Quality |
| Sinkou et al. 2017<sup>60</sup> | 4 | 5 | 4 | 3 | 3 | 5 | 4 | 4 | 32 (90%) High Quality |
| Stoner et al. 2022<sup>8</sup> | 5 | 5 | 4 | 5 | 5 | 5 | 5 | 5 | 39 (97.5 %) High Quality |
| Story et al. 2019<sup>9</sup> | 4 | 5 | 4 | 5 | 4 | 4 | 5 | 5 | 36 (90%) High Quality |
| Subbaraman et al. 2018<sup>70</sup> | 4 | 5 | 4 | 4 | 4 | 4 | 4 | 4 | 33 (82.5%) High Quality |
| Truong et al. 2022<sup>71</sup> | 5 | 5 | 5 | 5 | 4 | 3 | 4 | 4 | 35 (87.5%) High Quality |
| Wade et al. 2012<sup>61</sup> | 5 | 5 | 5 | 4 | 5 | 5 | 4 | 5 | 38 (95%) High Quality |
| Zúñiga et al. 2016<sup>62</sup> | 4 | 4 | 4 | 3 | 5 | 4 | 4 | 4 | 32 (80%) High Quality |

**Supplementary Figure 1:**
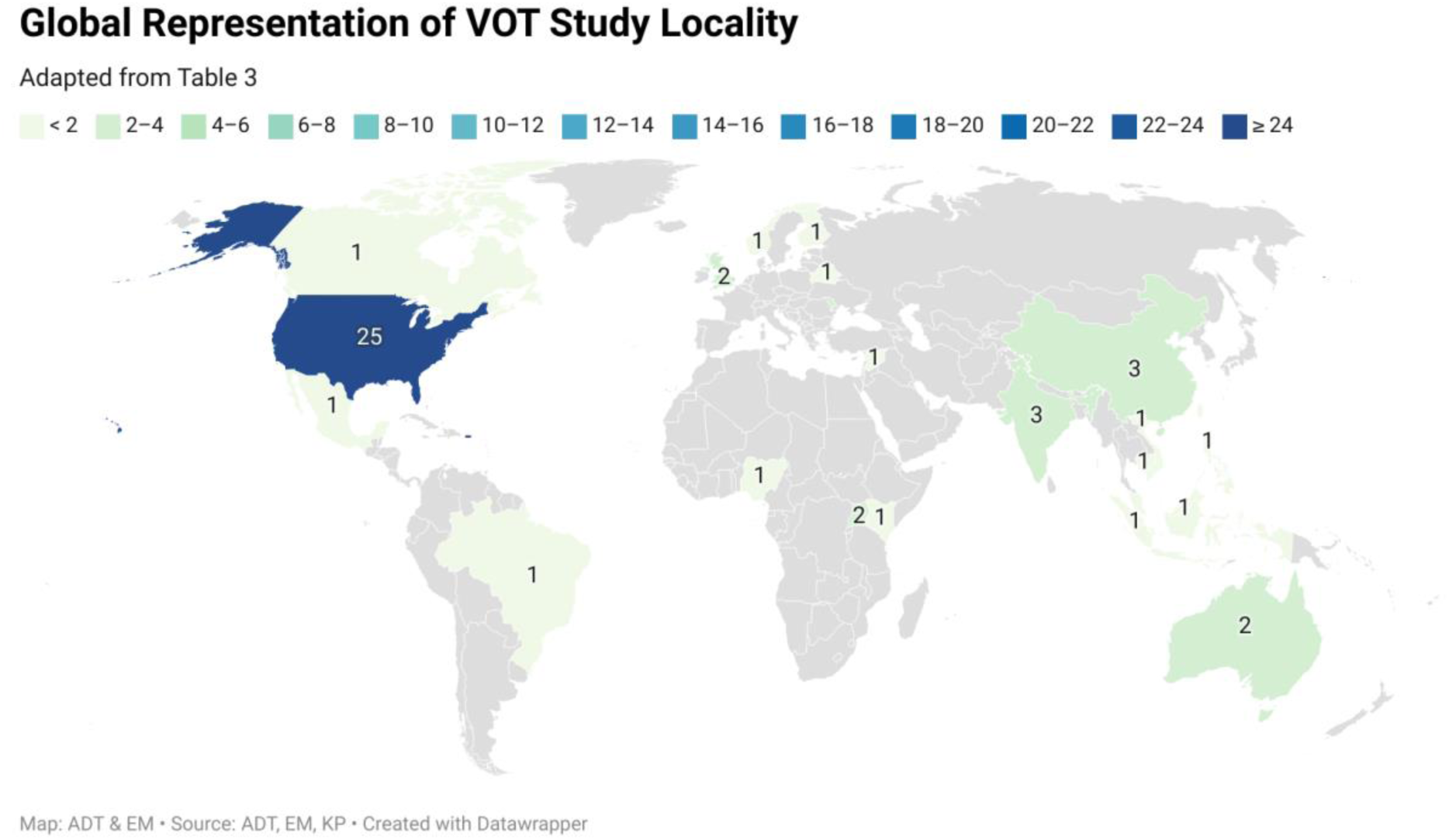
Geographical locations of study sites of VOT studies (n=55) *Legend: Due to country size or geopolitical recognition, Moldova (n=3, lower-middle income), Singapore (n=1, high income) and Taiwan (n=1, high income), are not visualised in the above map by the software used (Datawrapper, 2023). None of these three countries are high TB, HIV/TB, or MDR-TB burden countries.

